# Erythrocyte Sedimentation Rate in COVID-19 Infections

**DOI:** 10.1101/2020.06.25.20139881

**Authors:** Wei Zhang, Youshu Yuan, Shucheng Zhang, Can Jin, Linlin Wu, Hong Mei, Miao Chen, Zhixia Jiang, Zhixu He

**Author notes:** Equally Contributors (Wei Zhang, Youshu Yuan, Shucheng Zhang). Corresponding authors: Wei Zhang, MD: Institute: Department of Critical Care Medicine, Affiliated Hospital of Zunyi Medical, University; Address: 149 Dalian Road, Zunyi, Guizhou, 563000, China., Zhixu He, MD: Institute: Zunyi Medical University; Address: 6 Xuefu West Road, Xinpu New Area, Zunyi, Guizhou, 563003, China. (Email: Can Jin; Linlin Wu; Hong Mei; Miao Chen). (Youshu Yuan,). (Email: Shucheng Zhang; Zhixu He). (Email: Zhixia Jiang).

## Abstract

**Objectives:** To compare the clinical characteristics between the rapid cohort and the normal cohort of erythrocyte sedimentation rate (ESR) in COVID-19 infections, analyze the variables with significant differences, and explore the influencing factors of rapid ESR.

**Methods:** Selected a total of 80 patients with ESR detection during hospitalization were measured in 146 patients who received medical observation in concentrated isolation hospital in Guizhou province in China, collected and compared demographic information, epidemiological data, clinical symptoms, laboratory test data and CT image data during the observation between rapid cohort and normal group of ESR.

**Results:** By comparison, the proportion of male in the rapid cohort was higher than female. The average age was more than 35 years old, with a large age gap. The proportion of severe and critical patients was more than 26.53% (13/49). However, in the normal cohort the proportion of female was more than male, and the average age was about 8 years lower than the rapid cohort, and the age gap was smaller. The proportion of severe and critical patients was 12.90%, which was less than half of the rapid group. In the two groups, the proportion of clustered cases accounted for more than 50%, and the average number of patients in one family was more than 3. The most common clinical symptoms were cough, sputum, fever, sore throat and weakness of limbs. There were significant differences in ALT, γ-GT and C-reactive protein between the rapid and normal cohort (P<0.05), but no statistically significant in other indicators. Hemoglobin and C-reactive protein have a significant effect on erythrocyte sedimentation rate.

**Conclusions:** In this study, we found that ESR is related to Hemoglobin and C-reactive protein. (Funded by Science and Technology Department of Guizhou Province; Chinese ClinicalTrials.gov number, <u>ChiCTR2000033346. opens in new tab</u>.)

## Introduction

Hoffmann et al.^1^ have shown that the cell receptor of SARS-CoV-2 is angiotensin-converting enzyme 2 (ACE2), and SARS-CoV-2 is able to effectively bind ACE2 into cells and attack targeted organs that express ACE2, such as the lungs, heart, kidneys and gastrointestinal tract.^2^ Other studies have found that ACE2 is the receptor for SARS-CoV-2 to enter cells, which is highly expressed not only in alveolar epithelial cells and esophageal epithelial cells, but also in absorptive intestinal cells of ileum and colon. These results suggested both respiratory and digestive system became the potential route of infection.^3^ Furthermore, existing studies have shown that mutations or recombination in the regions of the spike protein and auxiliary protein of the SARS-CoV-2 genome are highly susceptible to occur.^4^ Toxicities during mutation may increase or decrease, and viral variability may have more severe consequences for humans. Therefore, we speculate that COVID-19 Spike proteins bind and destroy the ACE2 receptors of the mucosal epithelial cells of the respiratory tract or digestive tract, resulting in the loss of mucosal barrier function, thus opening up the door to the invasion of the body by the virus. After the virus invades further interacts with the body’s immune cells to produce inflammation storms, leading to a sharp increase in with a large number of inflammatory factors and acute-phase proteins such as C reactive proteins, cause uncontrolled systemic inflammatory reactions (SIRS) that may mediate autoimmune damage leading to a large number of immune complex deposition, further inducing the risk of autoimmune disease in the body.

Erythrocyte sedimentation rate (ESR) is an important index to reflect the immunological loss. However, COVID-19 as a highly infectious disease caused by SARS-CoV-2, which is not clear about the mechanism of how the virus interacts with the immune system after it invades the human body, except for the thorough research on the structure and genetic sequence of the virus. In the course of building the COVID-19 database, we found that the ESR obviously became rapid in many COVID-19 infections during hospitalization. In order to explore the mechanism of its occurrence, we designed this clinical trial.

## Methods

### Study design

This is a retrospective and single-center cohort study.

This retrospective, single-center cohort study was conducted on 80 COVID-19 infections who were admitted in Guizhou Provincial Staff Hospital in Guiyang, Guizhou, China from January 29, 2020 to April 30, 2020. The final follow-up data were collected on June 12, 2020. We obtained written informed consent from each participant. This study was approved by the Ethics Committee of Affiliated Hospital of Zunyi Medical University. This study was registered in Chinese Clinical Trial Registry Center. (CCTR number: ChiCTR 2000033346, registered 28 May 2020. URL: http://www.chictr.org.cn/edit.aspx?pid=53859&htm=4.)

### Patients

In this study, a total of 80 patients with ERS were selected from 146 patients who received medical observation in COVID-19 centralized isolation hospital in Guizhou province, among whom 49 patients had rapid ERS and 31 had normal ESR. Demographic information, epidemiological data, clinical symptoms, laboratory test data and CT imaging data of all patients were collected during observation.

### Detection methods of ESR

ESR is the sedimentation rate of red blood cells under certain conditions. The reason of acceleration is divided into physiological and pathological. According to the international standard method that is Westergren method of determination of erythrocyte sedimentation rate (ESR). the test instrument was automatic ESR analyzer which named Succeeder SD-100, and the blood samples were anticoagulant with 3.8% sodium citrate, the tube purchased from Zhejiang Gongdong medical device cooperative limited liability company, with a liquid solution of 1.6 ml. The normal value of ESR was 0 ∼ 15mm/h for males and 0 ∼ 20mm/h for females, indicating the faster rate of ESR outside the range

### Statistical methods

Use EXCEL software to conduct statistics on the data, and then import the collated data into SPSS 22.0 for analysis. The method of mean sequence is used to process the missing data. 2×2 cross table was used to measure the difference between the class variables in the two independent samples. When cases are more than 40 and the cell with the expected frequency less than 5 is not more than 20%, the chi-square test value is reasonable, otherwise the continuous correction chi-square value is read when the cell exceeds 20% and 1≤T<5. For the continuous quantitative variables in the two independent samples, the independent sample T test was used to compare the significance of the difference when the variables conforming to normal distribution. Non - parametric test was used to compare the non - normal distribution. Pearson correlation analysis was used to measure the degree of correlation between the two variables. The dependent variable ESR was a continuous numerical value, and multiple linear regression analysis was used to explore the factors and the degree of influence that work on the ESR among the multiple correlated variables.

### Study Definition

#### Classification

the standard for case classification in this study was based on COVID-19 diagnosis and treatment scheme, 5th to 7th edition from National Health Commission of the People’s Republic of China.

#### Epidemic focus

since 2019-nCoV first appeared in Wuhan, Hubei province, and then spread to the whole country and even the whole world, most of the existing studies regard Wuhan or other areas of Hubei province as the epidemic focus area.

#### Treatment cycle

The number of days of treatment cycle were actually counted in this study, which refers to the time from the first diagnosis to the definite detection show the patient was cured, excluding the observation time transferred to the centralized isolation hospital.

#### Cluster cases

The definition of cluster cases refers to the second edition of novel coronavirus pneumonia prevention and control program was released by the national health commission.

#### ESR acceleration window period

The duration of was the time from the onset date of the patient to the recovery period of the first ESR acceleration.

## Results

### CONSORT Flow Diagram (Figure 1)

In this research, the whole cohort encompassed 137 cases with COVID-19 infections including adults and children, as well as asymptomatic and confirmed COVID-19 infections. Eighty cases with ESR detection during hospitalization were divided into rapid ESR cohort and normal ESR cohort, we compared the epidemiological and clinical characteristics of the two cohorts.

**Figure 1.**
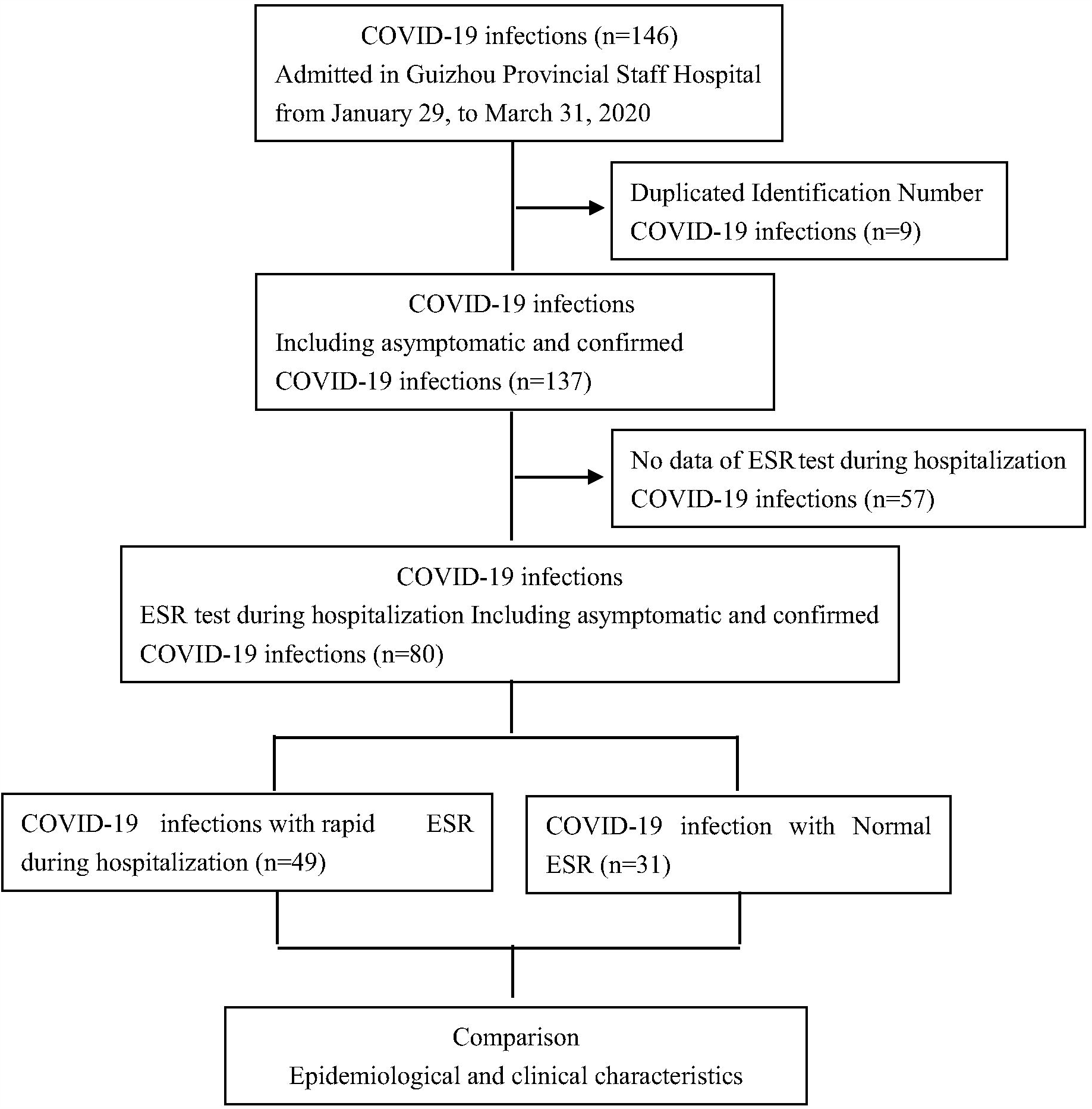
CONSORT Flow Diagram of This Study.

### Comparison of clinical features (Table 1)

A total of 80 patients with COVID-19 in this study, 31 (38.75%) had normal ERS and 49 (61.25%) had faster ERS. There were 26 males (53.06%) and 23 females (46.94%) in the faster group. The youngest patient was 6 years old and the oldest patient was 87 years old. The average age was 39.98 years, and the median age was 37 years (25th–75th percentile: 26∼53). In terms of classification, 5 (10.2%) patients were mild, 23 (46.94%) patients were general, 8 (18.37%) patients were severe, 5 (10.2%) patients were critical, and 7 (14.29%) patients were asymptomatic. Among them, 28 cases (57.14%) had a travel history in Wuhan or other areas of Hubei province. 36 cases (73.47%) were infected by cluster, with an average of 3.4 cases in one family. The median incubation period was 8 days (25th–75th percentile: 0∼16), and the incubation period of 17 patients was more than14 days. The average hospitalization period was 13.51 days. The most common clinical symptoms were fever in 18 cases (36.7%), cough in 19 cases (38.8%), expectoration in 9 cases (18.37%), weakness of limbs in 6 cases (12.2%). Other symptoms included runny nose, dizziness, pharyngeal pain, chills, shortness of breath et al. There were 5 patients with 1∼2 underlying diseases, including 3 patients with hypertension (10.2%) and 2 patients with diabetes and hypertension (4.1%).

There were 31cases in the normal group, including 11 males (35.48%) and 20 females (64.52%). Age range from 2 to 67 years old, the mean age was 32.16 years old, and the median age was 33 (25th–75th percentile: 20.5∼44) years old. The final diagnosis was 5 (16.13%) mild patients, 18 (58.06%) general patients, 3 (9.68%) severe patients, 1 (3.23%) critical patients, and 4 (12.90%) asymptomatic patients. 13 patients (41.94%) had a history of travel to the Hubei province. The 22 cases (70.97%) were clustered, and the average number of cases in one family was 3.6. The median incubation period was 4 days (25th–75th percentile: 0∼9.5), and the incubation period of 5 cases was more than14 days. The average hospitalization period was 13.13 days. The most common clinical symptoms were fever in 10 cases (32.26%), cough in 12 cases (38.71%), expectoration in 6 cases (19.35%), pharyngeal pain in 4 cases (12.90%). Other symptoms include weakness of limbs, dizziness, chest tightness, nasal obstruction, diarrhea, headache and shortness of breath et al. There was only one patient with primary hypertension.

**Table 1.** Baseline and Clinical Characteristics of Patient Population with COVID-19 (N=80)

|  | No.(61.25%)<br>Faster (n=49) | No.(38.75%)<br>Normal (n=31) | *p* value |
| --- | --- | --- | --- |
| **Sex** |  |  | 0.124 |
| Female | 23(46.94) | 20(64.52) |  |
| Male | 26(53.06) | 11(35.48) |  |
| **Age(y)(median [25th–75th percentile])** | 37(26~53) | 33(20.5~44) | 0.936 |
| Minimum | 6 | 2 |  |
| Maximum | 87 | 67 |  |
| Average | 39.98 | 32.16 |  |
| Mode | 30 | 36 |  |
| **Treatment Cycle** |  |  | 0.734 |
| Minimum | 3 | 3 |  |
| Maximum | 31 | 26 |  |
| Average | 13.51 | 13.45 |  |
| (days median [25th–75th percentile]) | 13(9~16) | 12(10~16.5) |  |
| **Epidemiology Investigation** |  |  |  |
| Epidemic Focus | 28(57.14) | 13(41.94) | 0.185 |
| Family Aggregation | 36 (73.47) | 22(70.97) | 0.807 |
| Average Familial Infection Number | 3.4 | 3.6 |  |
| **Initial symptom** |  |  |  |
| fever | 18(36.7) | 10(32.26) | 0.683 |
| cough | 19(38.8) | 12(38.71) | 0.995 |
| sputum | 9(18.37) | 6(19.35) | 0.912 |
| Sore throat | 2(4.1) | 4(12.90) | 0.2 |
| Limbs weakness | 6(12.2) | 3(9.68) | 1 |
| **underlying disease** |  |  | 0.472 |
| hypertension | 5(10.2) | 1(3.23) |  |
| diabetes | 2(4.1) | 0 |  |
| **Laboratory data on admission** |  |  |  |
| Hemoglobin |  |  | 0.100 |
| <110 | 9(18.37) | 3(9.68) |  |
| >160 | 0 | 1(3.23) |  |
| White blood cell count, $\times 10^9/L$ | | | 0.089 |
| <4 | 14(28.57) | 6(19.35) |  |
| Lymphocyte count, $\times 10^9/L$ | | | 0.065 |
| <1.1 | 10(20.4) | 5(16.13) |  |
| >3.2 | 1( 2.0) | 2(6.45) |  |
| Platelet count, $\times 10^9/L$ | | | 0.986 |
| <100 | 0 | 1(3.23) |  |
| >300 | 3(6.1) | 2(6.45) |  |
| C-response protein, mg/L |  |  | 0.000* |
| >10 | 6(12.2) |  |  |
| Neutrophils |  |  | 0.771 |
| <0.4 | 2(4.1) | 3(9.68) |  |
| >0.75 | 4(8.2) | 0 |  |
| **AST** |  |  | 0.429 |
| >40 | 8(16.3) | 3(9.68) |  |
| **ALT** |  |  | 0.049* |
| >40 | 13(26.5) | 6(19.35) |  |
| **BUN** |  |  | 0.730 |
| <2.9 | 18(36.7) | 12(38.71) |  |
| >7.5 | 2(4.1) | 0 |  |
| **LDH** |  |  | 0.715 |
| <109 | 10(20.4) | 3(9.68) |  |
| >245 | 3(6.1) | 0 |  |
| **γ-GT** |  |  | 0.007* |
| <3 | 1(2.0) | 0 |  |
| >50 | 20(40.8) | 6(19.3) |  |
| **CT Changes** |  |  | 0.995 |
| One side | 9(18.37) | 6(19.35) |  |
| Two side | 10(20.41) | 6(19.35) |  |

By comparison, the proportion of males in the faster group was higher than females. The average age was more than 35 years old, with a large age gap. The proportion of severe and critical patients was more than 26.53% (13/49) (Figure 3). The average age of the normal group was about 8 years lower than faster group, and the age gap was smaller. The proportion of severe and critical patients was 12.90%, which was less than half than the faster group. In the two groups, clustered cases accounted for more than half, and the average number of patients in one family was more than 3. The most common clinical symptoms were cough, sputum, fever, sore throat and weakness of limbs.

**Figure 2.**
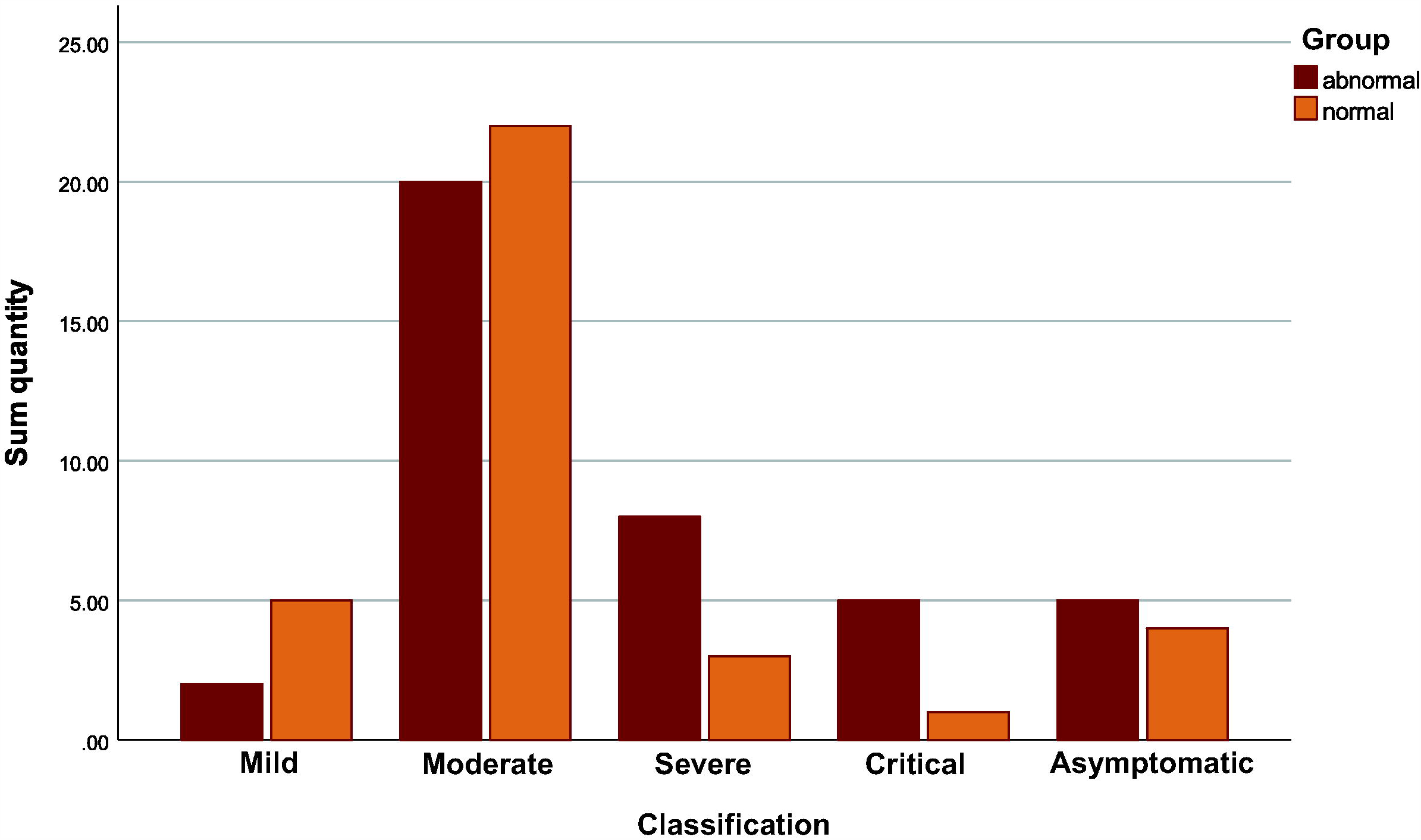
Distribution of Classification of Patients with Normal and Abnormal Sedimentation Rate.

**Figure 3.**
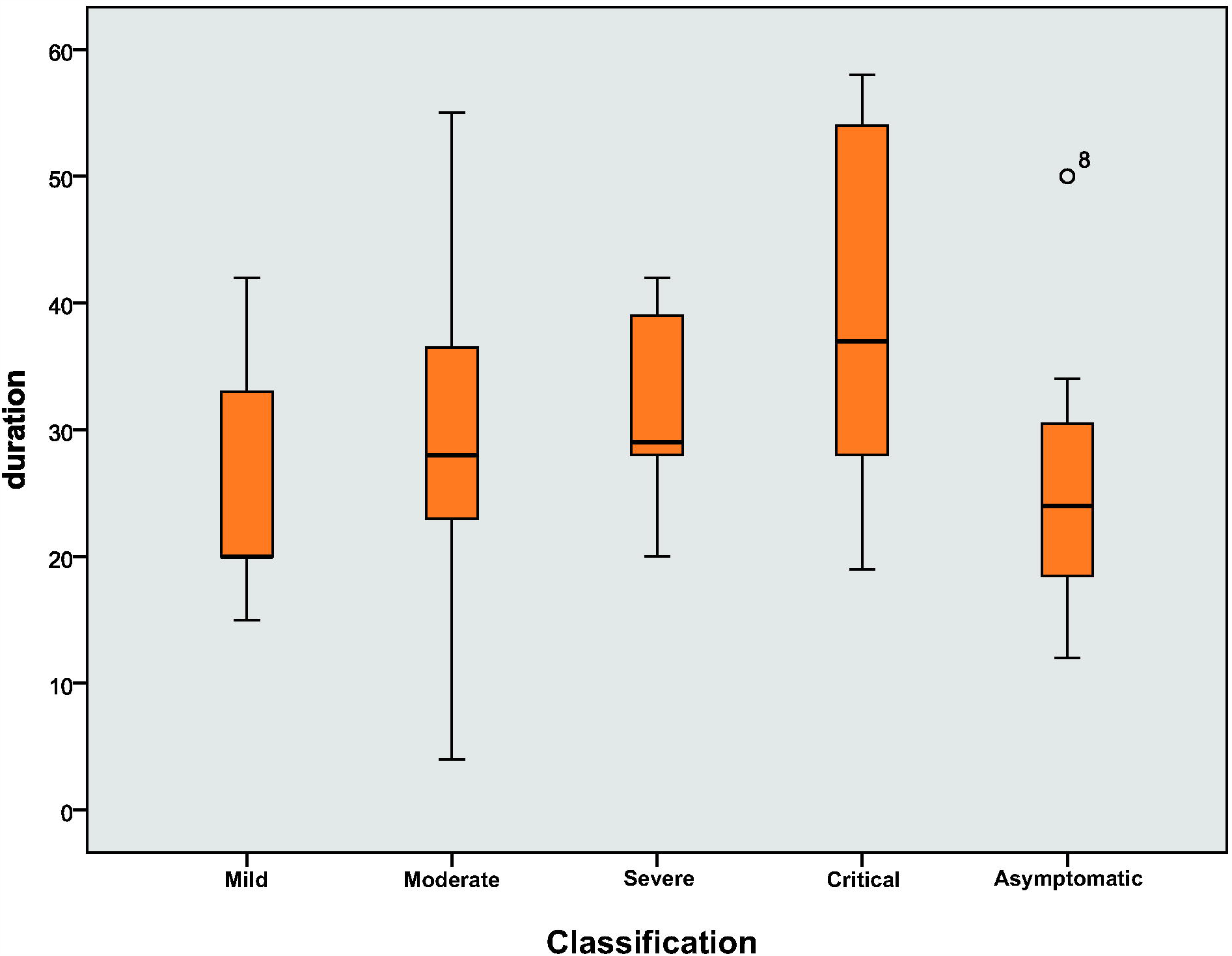
Comparison of Duration between Different Classifications in Rapid Sedimentation Rate.

### Comparison of laboratory data

9 patients (18.37%) had decreased hemoglobin in the faster group. 14 cases (28.57%) had decreased WBC count. 3 patients (6.1%) had increased platelets. Lymphocyte count was decreased in 6 cases (12.2%) and increased in 8 cases (16.3%). The absolute value of lymphocytes decreased in 10 cases (20.4%). Neutrophils were increased in 4 patients (8.2%). AST increased in 8 cases (16.3%). ALT was increased in 13 patients (26.5%). γ-GT was increased in 20 cases.CK was reduced in 1 case (2.0%). CK-MB was increased in 2 (4.1%) patients. Cr was decreased in 25 cases (51.02%). BUN was decreased in 18 cases (36.73%). LDH was reduced in 10 cases (20.41%). CRP was increased in 6 patients (12.24%).

Hemoglobin was decreased in 3 cases (9.68%) and increased in 1 case (3.23%) in the normal group. 6 cases (19.35%) had decreased WBC count. 2 patients (6.45%) had increased platelets. Lymphocyte count was decreased in 5 cases (16.13%) and increased in 3 cases (9.68%). Neutropenia was decreased in 3 patients (9.68%). AST was increased in 3 cases (9.68%). ALT was increased in 6 cases (19.35%). γ-GT had increased in 6 cases (19.35%). All patients with CK were normal. CK-MB was increased in 7 patients (22.58%). Cr was decreased in 19 cases (61.29%). 12 cases (38.71%) had BUN decreased. LDH was reduced in 3 cases (9.68%). CRP was all normal.

By comparison, the two groups showed the same trend in leucocyte, AST, ALT, CK-MB and Cr. Among them, there was a decreasing trend in the leukocyte and Cr. The expression of AST, ALT and CKMB increased. It is consistent with the existing studies that the total number of leukocytes in peripheral blood decreases in the early stage of the disease, and liver enzymes, and myozymes increased can be seen.^5^

### Comparison of chest CT examination

A total of 19 patients in the faster group developed lung lesions, including 9 patients with pulmonary one-side change and 10 patients with double-lung infection. In the normal group, a total of 12 patients developed pulmonary lesions, 6 patients had lobule changes in one side, and 6 patients were involved in both lungs.

### Results of difference analysis between two groups

By calculation, there was only exist significant difference of P<0.05 in ALT,γ-GT and C-reactive protein between the faster group and the normal group, but no statistical difference in other indicators. (Table 1)

### Analysis of influence factor to speed up ESR

#### The comparison of window period of faster ESR in different classification

As shown in figure 2, the average window period of faster ESR in asymptomatic was 44.14 days, and the median window period was 34 days (25th–75th percentile: 27.5∼53.5). The mean window period in mild patients was 38.6 days, and the median window period was 31 (25th–75th percentile: 31∼52) days. The average window period was 39.87 days in general patients, and the median window period was 34 days (25th–75th percentile: 25.5∼56). The mean window period in severe patients was 38.33 days, and the median window period was 28 days (25th–75th percentile: 19∼40). The mean window period in critical patients was 70.2 days, and the median window period was 64 (25th–75th percentile: 54∼94) days. For the normal and critical patients, the data were more discrete in the window period, and the window period in critical patients was significantly longer than other patients.

#### To explore the influence factors of faster ESR

##### Filter related variables

To analyze the correlation between ESR and other factors, such as sex, classification, age, hospitalization period, underlying disease, laboratory data, the results showed that age, underlying disease, hemoglobin, Cr, LDH and CRP six variables have significant correlation (Table 2), including underlying disease and Cr in Pearson correlation coefficient absolute value is less than 0.3, the correlation is very weak.

**Table 2.** The correlation between each variable and blood sedimentation rate.

| Correlated variable | Correlation<br>significance | Pearson's correlation<br>coefficient(\|r\|) |
| --- | --- | --- |
| ESR & age | 0.000 | 0.384** |
| ESR & Commodities | 0.035 | 0.236* |
| ESR & HB | 0.002 | 0.340** |
| ESR & Cr | 0.046 | 0.224* |
| ESR & LDH | 0.004 | 0.320* |
| ESR & C-response protein | 0.000 | 0.495** |

##### Explore the degree of Influence by Multiple Regression Analysis

The above six related variables were included into the independent variables of multiple linear regression analysis to explore whether there was a causal relationship between them and ESR. By calculation, the R^2^ between the independent variable and the dependent variable is 40.3%>30%, which means the degree that those six predictive variables such as age can explain the increase of ESR is 40.3%, which is in the acceptable range. The results showed that (Table 3), hemoglobin and C-reactive protein had a significant effect on ESR (Sig<0.05). The influence coefficient of hemoglobin on ESR was −0.311, indicating that hemoglobin had a slight negative influence on ESR. The influence coefficient of C-reactive protein on ESR was 1.683, which significantly positively affected the ESR, indicating that the higher the C-reactive protein value was, the higher the ESR would be.

**Table 3.** Analysis of influence factors of ESR.

| Possible influence factors | Influence<br>significance | Incidence (B) |
| --- | --- | --- |
| Age | 0.09 | 0.238 |
| Underlying disease | 0.528 | 3.063 |
| HB | 0.019 | - 0.311 |
| Cr | 0.216 | 0.167 |
| LDH | 0.193 | 0.074 |
| CRP | 0.001 | 1.683 |

## Discussion

### Age is correlated with ESR, but it is not necessarily cause and effect

This is a retrospective clinical study of patients with COVID-19 patients after were cured, who were discharged by Staff Hospital in Guizhou province. It was found that the proportion of severe and critical patients in the faster group was more than normal, and the average age was seven years older than normal group, which was consistent with the existing studies on the occurrence of severe and critical patients in the elderly more. Secondly, the youngest patient in the faster group was 6 years old, and the oldest patient was 87 years old, indicating a large age gap. The reason was considered to be the decline of immune function in the elderly and the immature immune system in children, both of which were susceptible to be infected by virus. It was speculated that due to the high plasticity of immune system in children, the probability of severe disease was lower than elderly patients. This study found that although there was a significant correlation between age and ESR, there was no causal relationship between two groups.

### ESR increased more rapidly in males than females, and cell receptor ACE2 was a key factor

A number of published literatures have reached inconsistent conclusions on the incidence and mortality of COVID-19 in different gender. The results of 44,672 cases reported by the Chinese center for disease control and prevention (CDC) showed that there was no statistical difference between male and female.^6^ However, the data of 4212 cases from South Korea showed that females were more susceptible to novel coronavirus than males.^7^ In this study, the faster ESR was more common in males. Studies on the pathogenesis of COVID-19 have shown that angiotensin converting enzyme 2 (ACE2) is the most critical molecule to control novel coronavirus infection. When SARS-CoV-2 virus contacts with respiratory tract, its spike protein binds to the cell surface receptor ACE2 and enters human lung cells to induce infection.^8^ Therefore, in this study, the faster ESR was mainly observed in males for the following reasons: first, ACE2 was a key molecule invaded by SARS-CoV-2 virus, which was positively regulated by androgen receptor, and its expression and distribution in male lung cells were higher than females. Second, the number of type II alveolar cells (AT2) expressing ACE2 in the lungs of male was significantly higher than female. Third, when male lung cells are attacked by the virus, the autoimmune response is weaker than female.^9,10^

### C-reactive protein can be used as a predictor factor after healing

Hemoglobin is a special protein that transports oxygen in red blood cells. Its movement in red blood cells must affect the sedimentation rate of red blood cells. C-reactive protein (CRP) is an acute inflammatory factor that plays a crucial role in the diagnosis and treatment of infectious diseases.^11^ In this study, C-reactive protein (CRP) significantly positively affected the erythrocyte sedimentation rate (ERS), that is the increase of CRP also result in significantly increased in ESR. The reason was that the infection with SARS-CoV-2 promoted the occurrence of inflammatory storms in patients with COVID-19. In order to enable the human body to have non-specific resistance to beat the virus, C-reactive protein played a positive role in the inflammatory response, which leads to its obvious rise. In addition, studies have demonstrated that continuous measurement of the inflammatory marker C-reactive protein can be used as a predictor factor in prognostic.^12^ In combination with this study, C-reactive protein has been found to have a positive effect on ESR, so it can predict that patients with COVID-19 will be susceptible to lupus erythematosus, rheumatoid arthritis and other connective tissue diseases after healing. The reasonable explanation was that the infection with novel coronavirus caused a significant increase in CRP, which leads to the increase in ESR, that could induce connective tissue disease.

### Research strengths and weaknesses

This study compared the difference between faster and normal in demographic characteristics, epidemiological history, physical examination and imaging. After demonstrating the variables with significant differences between the groups, multiple linear regression analysis was conducted to reveal the influencing factors and the degree in ESR, which was the first study to explore the influencing factors of faster ESR based on the clinical data of COVID-19 patients.

However, this study has several limitations to acknowledge. First, it was a retrospective study, we collected data prospectively from registries and medical records systems. Secondly, small sample size is a recognized limitation of data research. In view of the limited number of COVID-19 patients in Guizhou province and the difficulty in collecting continuous availability of patient clinical data, the sample size of this study is small. Therefore, it is suggested to expand the sample size with the data from surrounding provinces for verification at later stage.

## Conclusions

To sum up, novel coronavirus infection will lead to systemic inflammatory response and immune system dysfunction, and various systems of human were likely to have different degrees’ damage. However, due to individual differences in immune system, COVID-19 patients also have different progression or prognosis. C-reactive protein as an inflammatory marker and a reliable prognostic factor, has a significant positive effect on ESR, which can be used as a reference to predict the likelihood of encountering other diseases after recovery.

## Data Availability

I had full access to all of the data in this study and I take complete responsibility for the integrity of the data and the accuracy of the data analysis.

http://www.chictr.org.cn/edit.aspx?pid=53859&htm=4.

## Declaration

### Ethics approval and consent to participate

Written consent from the patients or their relatives was signed, although entirely anonymized data from which the individual cannot be identified. This study was approved by the Biomedical Ethics Committee of Affiliated Hospital of Zunyi Medical University.

### Consent to publish

Although entirely anonymized data and images from which the individual cannot be identified in this paper, we have access to the consent form from all participants. All authors have read and approved the manuscript version, and agree to submit for consideration for publication in the journal. We confirm that we have read the Journal’s position on issues involved in ethical publication and affirm that this report is consistent with those guidelines.

### Availability of data and materials

We stated that all the data and materials were true and available in the study.

### Authors’ contributions

Wei Zhang had full access to all data in the present study and accepts responsibility for data management and accuracy of the data analyses. Study concept and design: Wei Zhang, and Zhixu He. Acquisition and interpretation of data: Can Jin, Shucheng Zhang, and Youshu Yuan. Drafting of the manuscript: Youshu Yuan, Wei Zhang, and Shucheng Zhang. Critical revision of the manuscript for important intellectual content: Can Jin, Linlin Wu and Hong Mei. Administrative, technical, or material support: Miao Chen, Zhixia Jiang, and Shucheng Zhang. Study supervision: Wei Zhang and Zhixu He. All authors agree to submission of the final version of this manuscript. Wei Zhang is the study guarantor.

## Acknowledgements

The authors thank Ping Lu, Min Yao, Yang Hong, Jinhua Wu, et.al. gave us unselfish helps on the process of database construction of COVID-19 in this study.

